# T2 heterogeneity as an *in vivo* marker of microstructural integrity in medial temporal lobe subfields in ageing and mild cognitive impairment

**DOI:** 10.1101/2020.11.03.20225177

**Authors:** Alfie R. Wearn, Volkan Nurdal, Esther Saunders-Jennings, Michael J. Knight, Christopher R. Madan, Sean-James Fallon, Hanna K. Isotalus, Risto A. Kauppinen, Elizabeth J. Coulthard

**Affiliations:** Bristol Medical School, University of Bristol, Bristol, UK; School of Psychological Science, University of Bristol, Bristol, UK; Clinical Neurosciences, North Bristol NHS Trust, Bristol, UK; School of Psychology, University of Nottingham, Nottingham, UK; National Institute for Health Research Bristol Biomedical Research Centre, University Hospitals Bristol NHS Foundation Trust and University of Bristol, Bristol, UK; Faculty of Engineering, University of Bristol, Bristol, UK

**Keywords:** Magnetic resonance imaging, Alzheimer’s disease, Early diagnosis, Ageing, Hippocampal subfields, Medial Temporal Lobe

## Abstract

A better understanding of early brain changes that precede loss of independence in diseases like Alzheimer’s disease (AD) is critical for development of disease-modifying therapies. Quantitative MRI, such as T2 relaxometry, can identify microstructural changes relevant to early stages of pathology. Recent evidence suggests heterogeneity of T2 may be a more informative measure of early pathology than absolute T2. Here we test whether T2 markers of brain integrity precede the volume changes we know are present in established AD and whether such changes are most marked in medial temporal lobe (MTL) subfields known to be most affected early in AD. We show that T2 heterogeneity was greater in people with mild cognitive impairment (MCI; n=49) compared to healthy older controls (n=99) in all MTL subfields, but this increase was greatest in MTL cortices, and smallest in dentate gyrus. This reflects the spatio-temporal progression of neurodegeneration in AD. T2 heterogeneity in the entorhinal cortex also predicted cognitive decline over a year in people with MCI, where measures of volume or T2 in any other subfield or whole hippocampus could not. Increases in T2 heterogeneity in MTL cortices may reflect localised pathological change and may present as one of the earliest detectible brain changes prior to atrophy. Finally, we describe a mechanism by which memory, as measured by accuracy and reaction time on a paired associate learning task, deteriorates with age. Age-related memory deficits were explained in part by lower subfield volumes, which in turn were directly associated with greater T2 heterogeneity. We propose that tissue with high T2 heterogeneity represents extant tissue at risk of permanent damage but with the potential for therapeutic rescue. This has implications for early detection of neurodegenerative disease.

## INTRODUCTION

Accurate early diagnosis of Alzheimer’s disease (AD) is likely a necessity for development of disease-modifying therapies (Cummings et al., 2014, Alzheimer’s Association, 2015). Manifestation of cognitive symptoms, although required for clinical diagnosis, is a relatively late stage in the pathological process (Jack et al., 2010). Thus, clinical interventions after the appearance of cognitive deficits may be too late to restore brain health. A better understanding of the brain changes that precede loss of daily independence will help design early markers.

Structural and quantitative MRI show promise in their ability to identify changes in the brain that indicate early Alzheimer’s pathology. Identifying which people with mild cognitive impairment (MCI) will progress to AD dementia has been shown to be possible by measuring the volume of subfields within the medial temporal lobe (MTL) (Chételat et al., 2008, de Flores et al., 2015a, de Flores et al., 2015b, deToledo-Morrell et al., 2004, Apostolova et al., 2006, Apostolova et al., 2010). By its nature, a measurable change in volume is indicative of significant, and likely irreversible, atrophy. Smaller scale microstructural changes that occur prior to volume loss could help to identify patients in which such a treatment is to be optimally effective.

Recently, we demonstrated that the distribution width of T2 relaxation time (T2 heterogeneity) in the hippocampus predicted cognitive decline over a year in a group of people with MCI (Wearn et al., 2020a). We propose that T2 distribution widens because of different pathological hallmarks having opposing effects on T2, causing increased apparent heterogeneity without any change in absolute T2 (distribution midpoint). For example, non-haem iron, oligomers and plaques of β-amyloid (Aβ), and neurofibrillary tangles (NFTs) which build up around the MTL in early Alzheimer’s disease (Braak and Braak, 1991, Braak and Braak, 1995, Selkoe and Hardy, 2016, Smith et al., 2010) all cause T2 to decrease (Meadowcroft et al., 2015, House et al., 2008). In contrast, tissue alterations preceding necrosis cause cell membrane breakdown and oedema which increase the motility of water within a given region, subsequently causing T2 to increase (Laakso et al., 1996, Symms et al., 2004). These opposing factors necessitate examination of the heterogeneity of T2, rather than midpoint, for more accurate identification of microstructural impairment. This, we propose, is a reason for the lack of clear consensus from previous studies of T2 in AD, which exclusively look at absolute T2 (see Tang et al. (2018) for a review).

Our previous research focused on the T2 changes in the hippocampus as a whole (Wearn et al., 2020a). However, the hippocampus is not a uniform structure, rather, it comprises cytoarchitectonic subfields with distinct cellular structure, connectivity, functionality, and disease susceptibility (Duvernoy et al., 2013). NFTs first build up in the transentorhinal region of the MTL (Braak and Braak, 1995), which roughly corresponds to Brodmann area 35 (BA35), but also includes some of the lateral portion of entorhinal cortex (EC) (Xie et al., 2018). They then spread through EC, then to CA1, subiculum, other CA regions and finally dentate gyrus (DG) (Braak and Braak, 1995, Braak and Braak, 1991, Fukutani et al., 1995, Fukutani et al., 2000). Many of these changes occur even before symptom onset (Braak and Braak, 1991, Jack et al., 2003, Fukutani et al., 1995), highlighting their potential as prodromal markers. Reflective of histopathology, hippocampal volume loss due to AD is not uniform across the hippocampus – most atrophy is seen in CA1, though there is generally widespread volume loss in all or most subfields (La Joie et al., 2013, Mueller et al., 2010, Wolk et al., 2017, Kerchner et al., 2012, Sarazin et al., 2010, Frisoni et al., 2008, Frisoni et al., 2006). Volume loss is less severe in the hippocampus of people with MCI, often restricted to CA1 and, in many cases, the subiculum (Pluta et al., 2012, Mueller et al., 2010, Tang et al., 2014). For a review see de Flores et al. (2015a). This has even been shown in people who subjectively report cognitive decline but who have normal cognition as measured by standard cognitive tests (Perrotin et al., 2015).

Literature on T2 in MTL subfields is limited to very few *in vitro* studies (Huesgen et al., 1993, Antharam et al., 2012). As with the rest of the literature, these papers are focused on absolute T2. Antharam et al. (2012) do note that the distribution width of T2 within the main hippocampal subfields (CA4-DG and CA1-3) is wider in slices from AD patients than age-matched controls. However, they do not perform detailed analyses of the differences of distribution width between subfields. In a publication of pilot data from our group, Knight et al. (2019) concluded that T2 heterogeneity in MTL subfields can improve accuracy in distinguishing between healthy controls, those with MCI and Alzheimer’s disease patients. We know of no other studies that have explored quantitative T2 in subfields of the MTL *in vivo*.

The analyses in this paper are presented in two parts. In the first part, we test whether differences in T2 heterogeneity between subfields could distinguish healthy aging from MCI – important when considering T2 as a clinic tool to guide prognosis in MCI. We also analyse absolute T2 (distribution midpoint) to verify that it is heterogeneity and not absolute values that shift. Hypotheses:

1. The effect size of T2 heterogeneity increase in MCI (compared to healthy controls) will differ by subfield, reflective of the spatio-temporal progression of neurodegeneration in AD. Accordingly, T2 heterogeneity increases will be shown in the following pattern: BA35 > EC > CA > SUB > DG.
2. T2 heterogeneity in MTL subfields will better predict cognitive decline than in whole hippocampus in people with mild cognitive impairment. In the second part, we use path analysis to demonstrate the likely temporal sequence of neuroanatomical and behavioural changes in ageing. These are important to understand so that we track the right process at the right disease stage. Hypotheses:
3. Greater T2 heterogeneity indicates early damage that will lead to macroscopic structural change, and therefore will statistically mediate the relationship between age and volume of MTL subfields.
4. Subfield volume is indicative of macroscopic structural change, and therefore will mediate the relationship between T2 heterogeneity and cognitive performance.

## METHODS

The following methods are adapted from those presented by Wearn et al. (2020a).

The analyses in this paper combine data from two prospective longitudinal studies similar in cohort demographics and study design. No participants took part in both studies. Both studies are detailed in the following section. Where data collected are not identical between cohorts, we have normalised equivalent metrics within cohort and combined data after normalisation.

### Participants

Participants fulfilling the Petersen criteria (Albert et al., 2011) for diagnosis of MCI were recruited to both studies (Study 1: n=30, Study 2: n=29). Healthy older people (HC), with no history of memory problems or significant neurological disorders were recruited as controls to each study (Study 1: n=61; Study 2: n=56). All healthy controls had Montreal Cognitive Assessment (MoCA) > 26 (study 1) or Addenbrookes Cognitive Examination 3 (ACE-III) > 88 (study 2). 7 participants originally recruited as healthy controls in study 1 were found to have MoCA scores of <26, so were reclassified as MCI (given the high sensitivity and specificity of the MoCA for detecting MCI at this threshold; 90% and 100%, respectively (Nasreddine et al., 2005)).

Subjects for both studies were recruited from local GP surgeries and memory clinics in the Bristol area (having received MCI diagnoses or reported memory problems), Join Dementia Research, Avon and Wiltshire Mental Health Partnership’s Everyone Included system, an in-house database of volunteers, replies to poster adverts or through word of mouth. All patients provided informed written consent prior to testing as according to the Declaration of Helsinki. Ethical approval was given by Frenchay NHS Research Ethics Committee.

The current analyses included all participants who had volumetry and T2 relaxometry data for both hippocampal subfields, study 1 n=91 (50 HC, 30 MCI), study 2 n=66 (49 HC, 19 MCI). See tables 1 and 2 for demographic details. A total of 20 MCI participants were followed-up after one-year (10 from each study).

**Table 1.** Participants demographics. Cognitive score was calculated using MoCA (study 1) and ACE-III (study 2). Because of the different measures used between studies, each score was first normalised to each study’s respective HC group, before being pooled here. YOE = Years of Education, HC = Healthy Control, MCI = Mild Cognitive Impairment.

|  | HC | MCI | Total |
| --- | --- | --- | --- |
| N (male: female) | 99 (47:52) | 49 (27:22) | 148 (74:74) |
| Age (years) | 69.2 ± 8.55 | 72.2 ± 9.03 | 70.2 ± 8.79 |
| YOE | 15.8 ± 3.13 | 14.2 ± 2.81 | 15.3 ± 3.11 |
| Cognitive score<br>(normalised to HC) | .000 ± 1.00 | -3.75 ± 2.42 | -1.25 ± 2.39 |

**Table 2.** Summary statistics of key indirect paths. Data are not from independent tests, rather are from the same two multivariate models. Stringent correction for multiple comparisons across all variables is not warranted. All p-values below the uncorrected threshold for statistical significance (α) of .05 still remain below the threshold even after Bonferroni correction across two models (α=.025).

| Indirect path |  | Standardized estimate | P-value |
| --- | --- | --- | --- |
| Age → T2 heterogeneity → Volume |  |  |  |
|  | DG | -.072 | *.002 |
|  | CA | -.079 | *.002 |
|  | SUB | -.093 | *.002 |
|  | EC | -.103 | *.005 |
|  | BA35 | -.087 | *.009 |
| T2 heterogeneity → Volume → PAL |  |  |  |
| PAL Accuracy | DG | .063 | *.020 |
|  | CA | -.044 | .175 |
|  | SUB | -.032 | .177 |
|  | EC | .008 | .753 |
|  | BA35 | -.059 | *.013 |
| PAL Reaction Time | DG | -.050 | .130 |
|  | CA | .140 | *.002 |
|  | SUB | -.040 | .173 |
|  | EC | -.009 | .592 |
|  | BA35 | .021 | .220 |
| Age → T2 heterogeneity → Volume → PAL |  |  |  |
| PAL Accuracy | DG | .016 | *.013 |
|  | CA | -.016 | .135 |
|  | SUB | -.012 | .131 |
|  | EC | .003 | .749 |
|  | BA35 | -.024 | *.011 |
|  | Total path (all subfields) | -.006 | .183 |
| PAL Reaction Time | DG | -.013 | .096 |
|  | CA | .049 | *.002 |
|  | SUB | .056 | .137 |
|  | EC | -.004 | .585 |
|  | BA35 | .009 | .183 |
|  | Total path (all subfields) | .019 | .174 |

### Cognitive testing

Cognitive function was tested at baseline and follow-up using the MoCA in study 1 and the ACE-III in study 2.

Participants in both studies carried out the paired associates learning (PAL) task of the CANTAB toolbox which has shown high sensitivity to cognitive impairment and daily functioning in dementia (Égerházi et al., 2007).

### Imaging parameters

Scans for both studies were acquired on a Siemens Magnetom Skyra 3T system equipped with a parallel transmit body coil and a 32-channel head receiver array coil. The two studies used similar, but slightly different scanning protocols.

#### Study 1

This protocol has been previously described by Knight et al. (2019). The imaging protocol included a 3D T1-weighted whole-brain magnetization prepared rapid acquisition gradient-echo (MPRAGE) and 2D multi-contrast multi-spin-echo (CPMG).

##### MPRAGE

Coronal, whole-brain, repetition time (TR) 2200 ms, Echo Time (TE) 2.42 ms, Inversion time (TI) 900 ms, flip angle 9°, acquired resolution 0.68 x 0.68 x 1.60 mm, acquired matrix size 152 x 320 x 144, reconstructed resolution 0.34 x 0.34 x 1.60 mm (after two-fold interpolation in-plane by zero-filling in k-space), reconstructed matrix size 540 x 640 x 144, GRAPPA factor 2. Acquisition time: 5:25 min.

##### CPMG

Coronal, TR 4500 ms, TE 12 ms, number of echoes 10, echo spacing 12 ms, acquired resolution 0.68 x 0.68 x 1.7 mm inclusive of 15% slice gap, acquired matrix size 152 x 320, 34 slices, interleaved slice order, reconstructed resolution 0.34 x 0.34 x 1.7 mm (after two-fold interpolation in-plane by zero-filling in k-space, and inclusive of 15% slice gap), reconstructed matrix size 540 x 640, 34 slices, GRAPPA factor 2. Acquisition time: 11:07 min.

#### Study 2

This protocol has been previously described by Wearn et al. (2020a).

The imaging protocol included a 3D T1-weighted whole-brain MPRAGE and 2D multi-contrast turbo spin-echo (TSE).

##### MPRAGE

Sagittal, whole-brain, TR 2200 ms, TE 2.28 ms, TI 900 ms, flip angle 9°, FOV 220 x 220 x 179 mm, acquired resolution 0.86 x 0.86 x 0.86 mm, acquired matrix size 256 x 256 x 208. Acquisition time: 5:07 min.

##### Multi-contrast TSE

Coronal, TR 7500ms, number of echoes: 3, TE 9.1, 72 & 136 ms, acquired resolution 0.69 x 0.69 x 1.5 mm, reconstructed resolution 0.34 x 0.34 x 1.5 mm (after 2-fold interpolation in-plane by zero-filling in k-space, and inclusive of 15% slice gap), GRAPPA factor 2, FOV 220 x 220 x 34, acquired matrix size 270 x 320 x 58. Acquisition time: 5:09 min.

CPMG and TSE scans were not ‘whole-brain’, their coverage only extending approx. 1cm beyond anterior and posterior ends of the hippocampus. These scans were tilted such that the hippocampal body lay perpendicular to the slice acquisition plane.

The two distinct methods of measuring quantitative T2 (CPMG vs TSE) will give inherently different values for T2 midpoint and heterogeneity between studies (See supplementary information).

Relationships to variables such as age and cognitive score should be similar, given they are sensitive to the same tissue properties.

### Imaging analyses

All analyses were performed at CRICBristol in a Linux cluster environment. All analyses were carried out in single-subject native space.

CPMG and TSE scans were brain-extracted using FSL’s *bet2* on the first echo in the series (Smith, 2002). All extracted images were visually inspected for quality and rerun with different fractional intensity thresholds or gradient parameters where necessary. Fractional intensity threshold was typically set between 0.2-0.3. MPRAGE images were brain-extracted using *vbm8bet* (in-house script) and bias-field-corrected using FSL FAST (Zhang et al., 2001). T2 maps were created in MATLAB from multi-echo sequences by fitting logarithmic-space mono-exponential decay functions to each voxel series (overall summary of T2 calculation is shown in Knight et al. (2019)). The first echo of CPMG was always excluded. A sum-of-echoes image was created in order to have one structural image representing the entire multi-echo sequence. This image was used for segmentation.

Hippocampus was automatically masked using the Automatic Segmentation of Hippocampal Subfields (ASHS) software package (Yushkevich et al., 2015) (version: rev103, dated 12/06/2014; UPENN memory centre atlas dated 16/04/2014). ASHS has demonstrated high accuracy whilst minimising subjective rater bias, without the need for group blinding (Example output shown in Figure 1). CA1, CA2 and CA3 were pooled to create a total “CA” mask, given the small size of CA2 and CA3. Subfield masks were overlaid onto T2 maps, giving a value of T2 for each voxel within each subfield.

**Figure 1.**
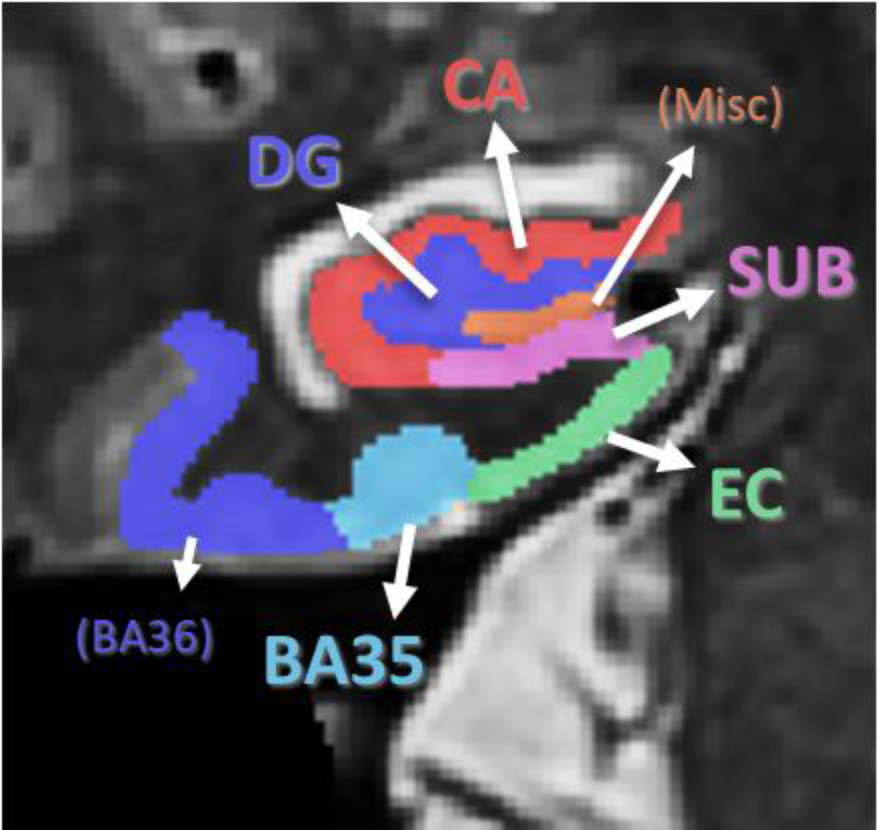
ASHS MTL mask example with subfields labelled. DG = Dentate Gyrus, CA = Cornu Ammonis 1-3, SUB=Subiculum, EC = Entorhinal Cortex, BA = Brodmann Area. Misc and BA36 were excluded from all analyses.

### Modelling T2 heterogeneity

T2 distribution histograms were modelled as loglogistic distributions within each subfield, as this was found to be the best fitting overall model in the whole hippocampus (Wearn et al., 2020a). Log-logistic distribution is defined as:

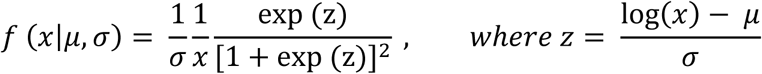

where μ and σ denote the log-median value (midpoint) and distribution shape (heterogeneity), respectively.

### Statistical analysis

Volumes, T2 parameters and general cognitive scores (MoCA / ACE-III) were pooled between studies after being converted into Z-scores for each study separately, with the left DG of healthy controls of each study as a reference point. In doing this, group, subfield, and hemisphere differences were maintained within each population.

We used a mixed model analysis, using the ‘*fitlme’* MATLAB function, to assess differences in T2 parameters between groups (HC, MCI), subfields (DG, CA, SUB, EC, BA35), hemispheres (left, right) and the effect of age. Full factorial models were created (assessing intercept and all possible interactions of aforementioned variables) with random effects of subject, subject*subfield interaction, and subject*hemisphere interaction. Each random effect significantly improved the model fit (as measured by AIC), without overparameterizing the model (confirmed by checking the hessian matrix and AIC). Age of the entire cohort was converted to a Z-score before being entered into the model. Models were created using Restricted Maximum Likelihood Estimation. Degrees of freedom were calculated using Satterthwaite approximation. A separate model was created for each of T2 midpoint (T2μ) and T2 heterogeneity (T2σ).

Between-group *post hoc* comparisons were assessed using independent-samples t-tests. Cohen’s *d* values for each comparison are reported as a measure of effect size. *Post hoc* effects of age in each subfield are assessed using linear regression.

Ability of volume and T2 in MTL subfields to predict cognitive decline was assessed using stepwise linear regression, with follow-up cognition as the dependent variable and baseline cognition and age as fixed covariates:

Follow-up Cognition

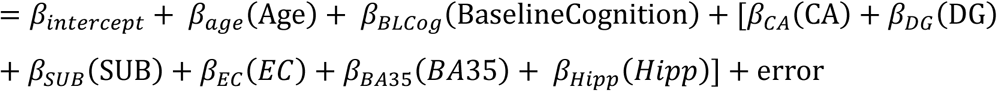

Parameters in square brackets represent those entered in a stepwise fashion. ‘Hipp’ represents total hippocampus. Z-scores for this analysis were calculated relative to each study’s MCI population only.

We assessed the relationship between age, subfield T2 heterogeneity, subfield volume (ICV-corrected) and memory using path analysis. Memory was assessed using two measures of performance on the PAL task – total accuracy and mean reaction time across all trials. We took a semi-supervised approach when defining the model paths. The direction of the arrows was predetermined by theory; T2 heterogeneity is caused by microstructural changes that precede significant atrophy. In this regard, volumes were not allowed to predict T2. Memory scores were not allowed to predict structural variables, and, for obvious reasons, no variable was allowed to predict age. All direct effects in this direction were modelled and compared. Initially the model was run with both PAL scores in a single model, feeding into a single latent variable representing overall PAL score. However, we observed poor factor loadings of each score onto the latent variable (<.60), so ran two separate models instead each with a single PAL outcome measure. Error terms were added to all variables except age and covariances and modification indices were calculated between all terms. All error term pairs within a structural measure (T2σ or volume) that had significant modification indices were allowed to covary, substantially improving overall model fit. We observed poor model fit when data was entered as Z-scores normalised to left DG of HCs of each study as described above. We therefore normalised each structural measure to only the respective HC population, such that the mean ± standard deviation for each structural measure within any given subfield was 0 ± 1. In other words, differences between subfields were not considered by the model. This improved model fit to acceptable levels and should be considered in interpretation of the data.

All reported p-values are two-tailed. Where possible we have used comprehensive models, to minimise the need for multiple comparison corrections. Balance tests were not carried out on demographic data for reasons detailed by Mutz et al. (2018). Data handling and storage was carried out using MathWorks MATLAB 2015a (with statistics and machine learning toolbox) and Microsoft Excel 2016. Mixed models were created and assessed in MathWorks MATLAB 2018a. Other statistical analyses were performed in IBM SPSS Statistics 24. Graphs were produced using GraphPad Prism v8.

## RESULTS

### Participant demographics

Demographic information for the cohort is displayed in Table 1. Our healthy control and MCI groups are closely matched for age and years of education. Details of cohorts in study 1 and study 2 are separately displayed in supplementary tables 1 and 2.

### Part 1: T2 changes in MCI

#### T2 heterogeneity

In addition to the overall effects of group (F(1,144)=17.7, p<.0001) and subfield (F(4,576)=262, p<.0001; explored further in supplementary information) and in line with hypothesis 1, the mixed model analysis revealed a significant group*subfield interaction (F(4,576)=5.01, p=.001). This effect did not vary according to hemisphere so a two-way ANOVA on pooled hemispheres was conducted with predicted values from the mixed model analysis, with pairwise group comparisons for each subfield (Figure 2). This test revealed a significantly greater T2 heterogeneity in the MCI group in all subfields but with varying effect sizes (DG: t=3.26, p_corr_ =.008, Cohen’s *d*=0.59; CA: t=3.79, p_corr_ =.001, Cohen’s *d*=0.68; SUB: t=5.50, p_corr_ <.0001, Cohen’s *d*=0.99; EC: t= 6.63, p_corr_ <.0001, Cohen’s *d*=1.22; BA35: t=5.26, p_corr_ <.0001, Cohen’s *d*=0.96, all adjusted using Bonferroni correction for multiple comparisons).

**Figure 2.**
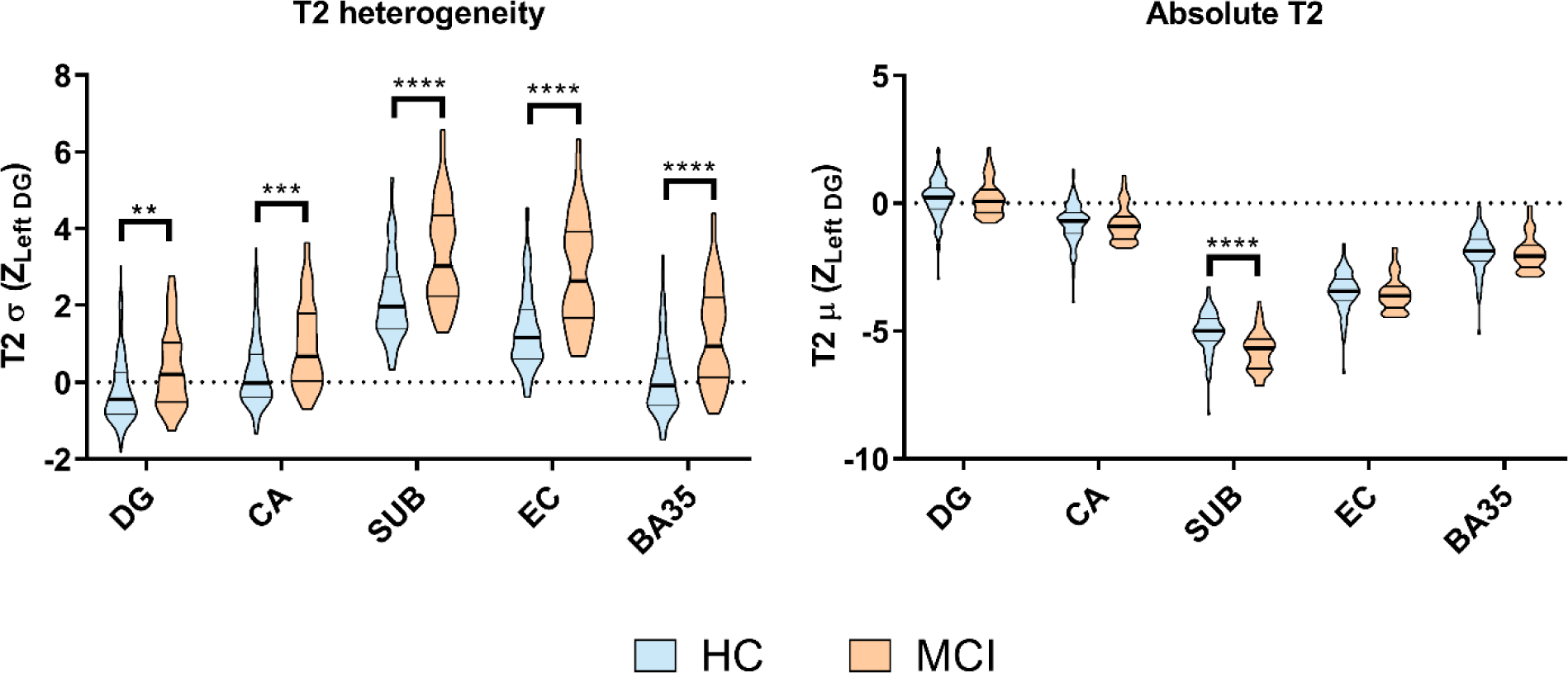
Subfield comparisons for T2 heterogeneity and absolute T2. Violin plots showing group & subfield differences (pooled across hemispheres & corrected for age) for absolute T2 and heterogeneity marginal means. HC = healthy older control; MCI = mild cognitive impairment. Stars represent p-values from post hoc two-way ANOVA tests to compare subfield groupwise differences. **p<.01; ***p<.001; ****p<.0001.

We also found a significant main effect of age (F(1,144)=37.6, p<.0001) and an interaction between subfield and age (F(4,576)=10.4, p<.0001). Older people had significantly increased T2 heterogeneity in all subfields. The association increased in strength in a stepwise fashion through DG (R^2^=.106, F(1,146)=17.3, p_corr_=.0003), CA (R^2^=.147, F(1,146)=27.5, p_corr_<.0001), SUB (R^2^=.159, F(1,146)=27.5, p_corr_<.0001), EC (R^2^=.185, F(1,146)=33.2, p_corr_<.0001) and BA35 (R^2^=.227, F(1,146)=42.8, p_corr_<.0001).

Finally, we observed no interactions between group and age (F(1,144)=1.74, p=.190) or group and hemisphere (F(1,144)=0.31, p=.579) nor were there any three- or four-way interactions for T2 heterogeneity.

#### Absolute T2

For comparison, we also explored subfield-specific changes in absolute T2. In contrast to T2 heterogeneity we observed no overall difference between HC and MCI groups on absolute T2 (F(1,144)=1.19, p=.278), but did see a substantial overall difference in absolute T2 between subfields (F(4,576)=974, p<.0001; explored further in supplementary information). We found a group*subfield interaction (F(4,576)=3.78, p=.005) driven by a subiculum-specific low T2μ in the MCI group (t=5.50, p_corr_<.0001, Cohen’s *d*=0.95). In no other subfield was there any difference between groups.

Although we observed no significant main effect of age on absolute T2 (F(1,144)=0.01, p=.907), the model did reveal a significant interaction between subfield and age (F(4,576)=2.92, p=.021). However, in no subfield was there any statistically significant association between age and absolute T2 (all subfields: p > .250).

We observed no interactions between group and age (F(1,144)=.005, p=.946) or group and hemisphere (F(1,144)=1.65, p=.201) nor were there any three- or four-way interactions for absolute T2.

### Predicting cognitive change over time

In order to identify whether MTL subfields could predict cognitive decline, we ran three stepwise linear regressions to predict cognitive score of people with MCI after one year. One model was ran for each MRI modality: volume, T2 heterogeneity, absolute T2, whilst accounting for baseline cognitive score and age. With no subfield values in the model, age and baseline cognition were unable to significantly predict follow-up cognitive score (R^2^=.092, F(2,19)=.859, p=.441).

Of all three modalities, only T2 heterogeneity could accuracy predict cognitive change over time. In this model, greater T2 heterogeneity within EC predicted poorer cognition after the year (R^2^=.406, F(3,19)=3.65, p=.035; β_EC_=-.665, p=.010).

EC volume was also entered as a significant individual predictor of greater cognitive decline in the volume model (β_EC_=.595, p=.030), however the overall model was not statistically significant (R^2^=.329, F(3,19)=2.62, p=.087). Similarly, absolute T2 in DG was selected as a significant predictor of cognitive score after one year, with higher values predicting greater cognitive decline (β_DG_=-.494, p=.044), however the overall absolute T2 model was not statistically significant (R^2^=.301, F(3,19)=2.30, p=.117).

### Part 2: Sequential relationships across brain structure and behaviour

We used path analysis to explore predicted relationships between age, T2 heterogeneity, subfield volume and cognition. We took a semi-supervised approach when defining the model paths. The direction of the arrows was predetermined by theory. Because T2 heterogeneity is likely caused by microstructural changes that precede significant atrophy, volumes were not allowed to predict T2 heterogeneity. Memory scores were outcome variables, so were not allowed to predict structural variables, and no variable was allowed to predict age. All direct effects in this direction were modelled and compared. Two separate models were created, one for each of the PAL output measures (See Methods). After applying covariance structures to error terms between subfields, model fit was good with various fit parameters falling within an acceptable range (*χ*^*2*^=21.8, df = 20, p=.350; C_min_/DF=1.09; GFI=.964, AGFI=.861; CFI=.998; RMSEA=.031 ± 90% CI [<.001-.096]; p_close=_.615). Model fit was identical between the two models. Covariance matrices can be found in supplementary information (Supplementary Tables 3 & 4).

The final path analysis models (Figure 4) reveal the following relationships, for which statistics are shown in Table 2. Age is a significant positive predictor of T2 heterogeneity in all MTL subfields. In turn, T2 heterogeneity within each subfield is a significant negative predictor of subfield volume. The only significant direct effects of age on subfield volume are seen with DG and CA (where greater age predicts smaller volumes), however, significant indirect effects are seen between age and subfield volume within all subfields (Table 2). T2 heterogeneity therefore mediates the relationship between age and volume.

**Figure 3.**
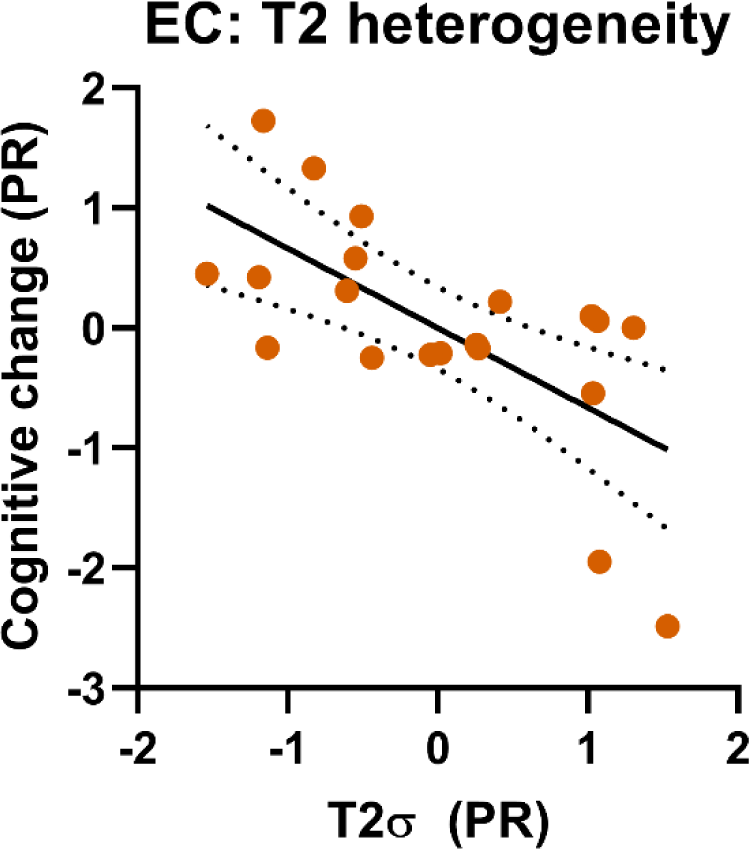
Subfield structure predicting cognitive decline in people with MCI. Partial residual (PR) plot of significant model from stepwise linear regression. Graph shows linear regression lines (solid lines) ± 95% confidence intervals (dotted lines). The only significantly predictive model created was that involving T2 heterogeneity within entorhinal cortex (EC).

**Figure 4.**
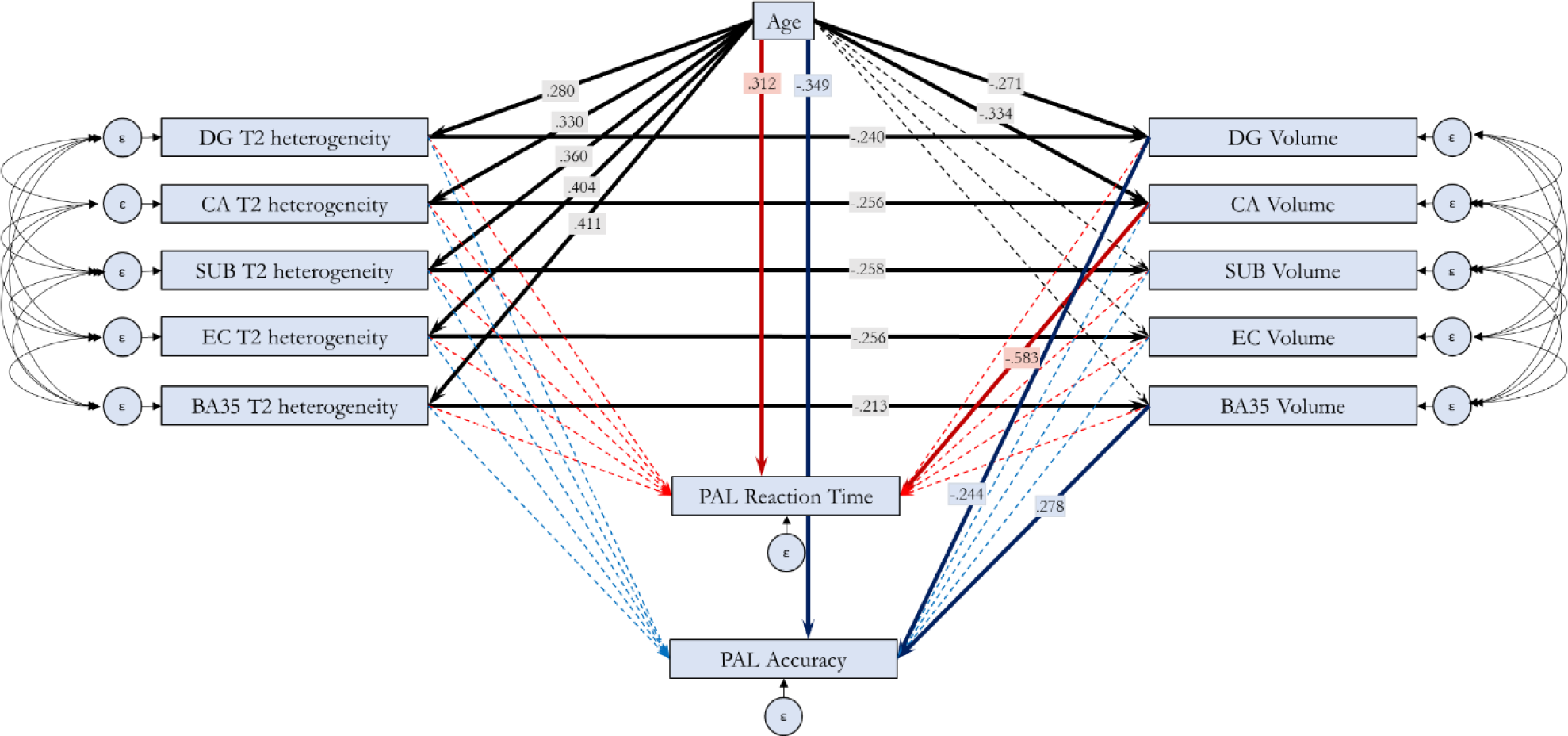
Path analysis showing the relationship between Age, T2 heterogeneity, volume and memory in MTL subfields in healthy older controls. Bold arrows represent statistically significant relationships (p<.05), with standardized B values indicated in overlaid boxes. Two models were run, each assessing one outcome measure of the PAL task. Black lines represent paths shared between the models. Unique paths to each model are shown in red (PAL reaction time as dependent variable) and blue (PAL Total Accuracy as dependent variable). Curves lines represent error term covariances defined in the model. All subfield volumes were normalised to ICV prior to entering into the model.

No significant direct effects were observed between T2 heterogeneity and either PAL score (Table 2). However, a direct negative effect of CA volume on PAL mean reaction time was observed, as well as a direct positive effect of BA35 volume on PAL accuracy. In contrast, we also see that DG volume has a direct negative relationship with PAL accuracy. The indirect relationship between T2 heterogeneity for each of these respective subfields and each respective PAL score is statistically significant (Table 2), indicating a role of volume as a mediator between T2 heterogeneity and cognition. Interestingly, analysis of indirect paths also reveals a marginally significant positive relationship between DG T2 heterogeneity and PAL reaction time.

Finally, outside of MTL subfield structure and microstructure, we also observe direct effects of age on PAL scores, with greater age predicting poorer scores on both measures (Table 2).

This all serves to show a clear pathway of the effect of age on cognition, first through its increasing of T2 heterogeneity in subfields, which in turn reduces the volume of each subfield, which then, in a subfield-specific manner, reduces cognitive functionality. The full indirect pathway from age, through T2 heterogeneity, to volume and finally to PAL score is statistically significant for pathways through DG and BA35 for predicting PAL accuracy, and through CA for predicting PAL reaction time (Table 2). Outside any MTL subfield paths, there remains a direct effect of age on PAL score, whereby greater age predicts poorer performance (lower accuracy, longer reaction times), suggesting that age affects PAL performance through non-hippocampal as well as hippocampal pathways.

We ran the models again on the MCI group data to explore any differences in path structure to the HC models. We observed no major changes between the models other than the loss of the direct relationship between age and PAL scores. This will not be discussed further here as it is not the focus of this study, but the model path diagram can be seen in supplementary information.

## DISCUSSION

Here we have shown that T2 heterogeneity in the MTL increases with cognitive impairment in a subfield-dependent manner in line with progression patterns of Alzheimer’s pathology. We also show that T2 heterogeneity in the entorhinal cortex predicts cognitive decline in people with MCI better than volume or T2 changes in any other subfield. Furthermore, we describe a mechanism by which cognitive ability deteriorates with age through direct effects on T2 heterogeneity, which lead to the changes in volume which in turn lead to cognitive decline. These findings are discussed in detail below.

### Subfield-specific differences in T2 heterogeneity in MCI and with age

T2 heterogeneity was greater in MCI across all subfields of the MTL, however the magnitude of this effect differed between subfields. The smallest increase was seen in DG, and the largest in EC and BA35. This is in line with literature on the timing of deposition of NFTs (Braak and Braak, 1991, Braak and Braak, 1995) throughout the course of AD, whereby the transentorhinal region (comprising BA35 and lateral EC) is affected first, and DG pathology is only detectible later on. DG is also one of the last MTL regions to exhibit volume loss (Daugherty et al., 2015). This accordance between the pattern of T2 heterogeneity differences and AD neuropathological progression supports T2 heterogeneity as a means to detect pathologically relevant change before the onset of dementia.

This is in contrast to absolute T2, which is no different between healthy controls and people with MCI in any subfield other than subiculum (where it is decreased in MCI). The effect may be explained by an increase in iron and compact amyloid plaques, which have been seen to be increased in SUB and CA1 regions compared to DG and CA3 in a mouse model of AD (Reilly et al., 2003). Even though we cannot directly show the prevalence of AD pathology in our MCI sample, these results are in line with this group being at higher risk for displaying AD pathology.

Our findings support our proposal (Knight et al., 2019, Wearn et al., 2020a) that T2 heterogeneity defines hippocampal integrity at the subfield level better than absolute T2. In further support of this model, two studies of absolute T2 in hippocampal subfields *in vitro* (Huesgen et al., 1993, Antharam et al., 2012) note a lack of any consistent pattern between healthy control and Alzheimer’s slices in any subfield.

### Clinical Utility of T2 heterogeneity in MTL subfields

T2 heterogeneity in EC is the best predictor of cognitive decline in our MCI cohort, outperforming any predictive ability of volume or absolute T2 in whole hippocampus or MTL subfields, as well as whole hippocampal T2 heterogeneity. This may enable identification of those who have MCI due to incipient Alzheimer’s disease as opposed to other causes, though further testing is required to confirm this.

The use of T2 heterogeneity is highly translatable to clinical settings. MRI is standard practice in improving the accuracy of a diagnosis of AD. The MRI scan necessary to calculate quantitative T2 can be completed within a few additional minutes of a standard clinical MRI (a multi-echo T2 sequence of sufficient resolution is all that is required). The same high-resolution scan can be used to automatically segment subfields of the MTL e.g. using ASHS (Yushkevich et al., 2015). The use of MRI in people with dementia can sometimes be tricky due exacerbated feelings of claustrophobia and confusion. However, this MRI protocol would be most useful in prodromal stages of the disease, before significant MTL volume loss and symptom severity, minimizing these complications. A cheaper theoretical screening test with high sensitivity for detecting pathology such as has been shown with tests of accelerated forgetting on word list tasks (Wearn et al., 2020b, Weston et al., 2018) or a blood test (Toombs and Zetterberg, 2020) may identify individuals who should qualify for an MRI scan sensitive to very early pathological hallmarks.

Although we have focussed on T2 changes due to Alzheimer’s pathology, T2 heterogeneity is a novel measure which could be applied to any neurological disease characterised by microstructural changes, including other dementias, acute stroke (as demonstrated by Norton et al. (2017)), epilepsy or schizophrenia. Future research could utilise T2 heterogeneity to easily probe microstructural abnormalities. This study builds on our past analyses of T2 heterogeneity by highlighting how it can reveal structural changes on an even finer scale, indicating its usefulness in disorders where brain damage is highly localised.

### Subfield-specific contributions to memory

We aimed to better understand the relationship between T2 heterogeneity, volume, age and cognitive ability in cognitively healthy older people. We employed the basic assumption that T2 heterogeneity would lead to changes in volume, rather than the other way around, which is supported by the strength of this model. To summarise, our path analysis reveals the following pattern of relationships. T2 heterogeneity mediates the negative relationship between age and volume. Volume in turn mediates negative relationship between T2 heterogeneity and memory, in a subfield-specific manner. In line with hypotheses 3 and 4, increased T2 heterogeneity therefore seems to represent a state of structural damage which may give rise to the volumetric changes which have the most profound impact on memory and cognition. Interestingly, we do still find a direct association between age and CA and DG volume, indicating only partial mediation by T2 heterogeneity in these regions. This suggests that there are other age-mediated volumetric changes which are not accurately reflected by changes in T2 heterogeneity.

Our results reveal subfield-specific associations with memory scores unlike previous similar models that have looked at whole hippocampus only (Rodrigue et al., 2012). Longer reaction times were associated with a subtle damage to the CA region, whereas poor accuracy was associated with low BA35 integrity but a more intact DG. This latter result was surprising but may suggest that an impaired BA35 *relative to* DG is indicative of a network prone to specific deficits in total accuracy. This pattern reflects changes seen in early AD, as the transentorhinal cortex is the first region to display NFT deposition, and DG is the last of all regions tested (Braak and Braak, 1995, Braak and Braak, 1991, Xie et al., 2018). In other words, patterns of very subtle neurodegeneration akin to those found in the earliest stages of AD may give rise to specific deficits in total accuracy scores on the CANTAB PAL.

### Limitations

The main limitation is the lack of biomarker status availability for people with MCI in this study (assessed either from Positron Emission Tomography or CSF analysis). Therefore, we cannot be certain as to the exact proportion of those who have incipient dementia. We suspect that T2 heterogeneity in entorhinal cortex would identify those who do have incipient Alzheimer’s disease, as EC is one of the earliest sites of pathological change. This is supported by the ability of EC T2 heterogeneity in predicting cognitive decline. However, co-pathologies are almost certainly present in the overall cohort which may include other dementias or undiagnosed microbleeds.

Additionally, both studies utilised different scanning sequences for assessing quantitative T2, giving inherently different absolute T2 values. For this reason, we were careful to normalise results from each cohort before combining them. Although a potential confounding factor, it is encouraging that the effects we see are not specific to a sequence. Rather, we provide evidence that the different sequences are sensitive to the same physiological changes. This supports the translatability of this measure to clinical settings, where available sequences for measuring quantitative T2 may vary between sites.

## Conclusions

The analyses presented in this paper comprise the first detailed exploration of quantitative T2 across subfields of the medial temporal lobe in older people with and without cognitive impairment. We support previous evidence that absolute T2 is not a sensitive marker of early pathology, rather, heterogeneity of T2 is much more sensitive to early pathological change and differs between subfields in a manner which reflects the order of NFT deposition in prodromal AD. We provide evidence that although T2 heterogeneity increases with age in all subfields, the degree to which this occurs is subfield-dependent and is strongest in MTL cortical regions (EC and BA35). In contrast, we do not see systematic evidence of a relationship between age and absolute T2. Using path analysis we describe a pathway through which cognition is significantly affected by age through direct effects on T2 heterogeneity in cognitively healthy older people, which in turn has direct effects on volume which lead to changes in cognition, supporting the idea that T2 changes precede and lead to volumetric changes. Finally, we show that the single best predictor of cognitive decline in people with MCI is greater T2 heterogeneity within entorhinal cortex, outperforming both T2 midpoint and, critically, volume of any subfield.

## Data Availability

The datasets used during the current study are available from the corresponding author on reasonable request.

## Declarations

### Ethics approval and consent to participate

All patients provided informed written consent prior to testing. Ethical approval was given by Frenchay NHS Research Ethics Committee.

### Consent for publication

Not applicable

### Competing interests

We declare that none of the authors have competing financial or non-financial interests.

### Funding

This research was funded in part by the Wellcome Trust [Grant number 109067/Z/15/AI]. For the purpose of open access, the author has applied a CC BY public copyright licence to any Author Accepted Manuscript version arising from this submission. This study also was funded by Alzheimer’s Research UK and BRACE.

### Authors’ contributions

This manuscript was written and cognitive testing protocol designed by A.W. and E.C. Imaging protocols were set up by R.K and M.K. M.K. also set up imaging analysis pipelines which were later managed by A.W. Cognitive testing, scoring and imaging was carried out by A.W., V.N., E.S-J., H.I.. Statistical analyses were performed by A.W., C.M., and S.J.-F.

## Acknowledgments

The authors wish to thank Join Dementia Research and the Avon & Wiltshire Mental Health Partnership for their assistance with participant recruitment. We also wish to thank those who have helped collect data for the projects (Serena Dillon, Demitra Tsivos, Emma Hadley, Ellen Gaaikema, Lucy Adams, Candida Stainer, Ben Kershaw & Bryony McCann), Aileen Wilson for her help conducting MRI scans, and all the volunteers who gave up their time to take part in our studies.

## Notes

### Competing Interest Statement

The authors have declared no competing interest.

