## Supplementary Information for "T2 heterogeneity as an *in vivo* marker of microstructural integrity in medial temporal lobe subfields in ageing and mild cognitive impairment"

### Individual study cohort information

Demographic information, neuropsychological test data and data from MRI summaries for study 1 and study 2 can be found in Supplementary Tables 1 and 2, respectively.

Supplementary Table 1 | Study 1 cohort information.

Demographic, neuropsychology, and MRI structural measure info for study 1 cohort. Data show mean ± standard deviation. Some neuropsychological measures are missing for a small number of participants. The numbers, and which groups they belong to, is indicated in the leftmost column. All volumes shown have been corrected for intracranial volume. HC = Healthy Control; MCI = Mild Cognitive Impairment; YOE = years of education; MoCA = Montreal Cognitive Assessment;; PAL = CANTAB Paired Associate Learning

| **Demographics** | HC | MCI |
| --- | --- | --- |
| N (male: female) | 50 (21:29) | 30 (14:16) |
| Age (years) | 67.7 ± 9.03 | 70.7 ± 8.55 |
| YOE | 15.6 ± 2.66 | 14.5 ± 2.70 |
| **Neuropsychological testing** |  |  |
| MoCA (/30) | 28.0 ± 1.32 | 23.0 ± 2.83 |
| PAL (-2 MCI) |  |  |
| Accuracy | .706 ± .131 | .534 ± .145 |
| Mean Reaction Time (ms) | 2120 ± 488 | 2960 ± 1490 |
| **MRI structural measures** |  |  |
| Right DG |  |  |
| T2σ | 0.09 ± 0.02 | 0.10 ± 0.02 |
| T2μ | 4.62 ± 0.03 | 4.62 ± 0.02 |
| Volume | 0.56 ± 0.07 | 0.50 ± 0.08 |
| Left DG |  |  |
| T2σ | 0.09 ± 0.02 | 0.10 ± 0.02 |
| T2μ | 4.62 ± 0.03 | 4.62 ± 0.02 |
| Volume | 0.54 ± 0.07 | 0.49 ± 0.08 |
| Right CA |  |  |
| T2σ | 0.09 ± 0.20 | 0.10 ± 0.02 |
| T2μ | 4.59 ± 0.03 | 4.59 ± 0.03 |
| Volume | 0.93 ± 0.13 | 0.80 ± 0.16 |
| Left CA |  |  |
| T2σ | 0.09 ± 0.20 | 0.10 ± 0.02 |
| T2μ | 4.60 ± 0.03 | 4.59 ± 0.02 |
| Volume | 0.91 ± 0.12 | 0.80 ± 0.15 |
| Right SUB |  |  |
| T2σ | 0.11 ± 0.02 | 0.13 ± 0.03 |
| T2μ | 4.48 ± 0.04 | 4.46 ± 0.03 |
| Volume | 0.24 ± 0.03 | 0.22 ± 0.04 |
| Left SUB |  |  |
| T2σ | 0.11 ± 0.02 | 0.13 ± 0.02 |
| T2μ | 4.49 ± 0.04 | 4.46 ± 0.04 |
| Volume | 0.25 ± 0.04 | 0.23 ± 0.05 |
| Right EC |  |  |
| T2σ | 0.10 ± 0.02 | 0.12 ± 0.03 |
| T2μ | 4.53 ± 0.04 | 4.53 ± 0.04 |
| Volume | 0.27 ± 0.05 | 0.24 ± 0.05 |
| Left EC |  |  |
| T2σ | 0.10 ± 0.02 | 0.12 ± 0.02 |
| T2μ | 4.52 ± 0.04 | 4.52 ± 0.04 |
| Volume | 0.31 ± 0.04 | 0.29 ± 0.07 |
| Right BA35 |  |  |
| T2σ | 0.09 ± 0.02 | 0.10 ± 0.02 |
| T2μ | 4.56 ± 0.04 | 4.57 ± 0.04 |
| Volume | 0.27 ± 0.06 | 0.25 ± 0.06 |
| Left BA35 |  |  |
| T2σ | 0.09 ± 0.02 | 0.10 ± 0.02 |
| T2μ | 4.55 ± 0.03 | 4.54 ± 0.03 |
| Volume | 0.26 ± 0.05 | 0.25 ± 0.07 |

Supplementary Table 2 | Study 2 cohort information.

Demographic, neuropsychology, and MRI structural measure info for study 2 cohort. Data show mean ± standard deviation. Some neuropsychological measures are missing for a small number of participants. The numbers, and which groups they belong to, is indicated in the leftmost column. All volumes shown have been corrected for intracranial volume. HC = Healthy Control; MCI = Mild Cognitive Impairment; YOE = years of education; ACE-III = Addenbrookes Cognitive Examination-III; PAL = CANTAB Paired Associate Learning.

| **Demographics** | HC | MCI |
| --- | --- | --- |
| N (male: female) | 49 (26:23) | 19 (13:6) |
| Age | 70.8 ± 7.79 | 74.5 ± 9.49 |
| YOE | 16.0 ± 3.57 | 13.7 ± 3.00 |
| **Neuropsychological testing** |  |  |
| ACE-III /100 | 94.9 ± 3.17 | 80.2 ± 6.62 |
| PAL (-2 HC, -4 MCI) |  |  |
| Accuracy | .695 ± .113 | .493 ± .184 |
| Mean Reaction Time (ms) | 2280 ± 485 | 3880 ± 2350 |
| **MRI structural measures** |  |  |
| Right DG |  |  |
| T2σ | 0.11 ± 0.01 | 0.11 ± 0.01 |
| T2μ | 4.70 ± 0.03 | 4.72 ± 0.03 |
| Volume | 0.48 ± 0.06 | 0.42 ± 0.08 |
| Left DG |  |  |
| T2σ | 0.12 ± 0.01 | 0.12 ± 0.01 |
| T2μ | 4.69 ± 0.03 | 4.69 ± 0.03 |
| Volume | 0.46 ± 0.06 | 0.39 ± 0.09 |
| Right CA |  |  |
| T2σ | 0.12 ± 0.01 | 0.13 ± 0.01 |
| T2μ | 4.67 ± 0.03 | 4.67 ± 0.03 |
| Volume | 0.79 ± 0.09 | 0.65 ± 0.15 |
| Left CA |  |  |
| T2σ | 0.12 ± 0.01 | 0.13 ± 0.01 |
| T2μ | 4.67 ± 0.03 | 4.67 ± 0.04 |
| Volume | 0.79 ± 0.10 | 0.62 ± 0.15 |
| Right SUB |  |  |
| T2σ | 0.15 ± 0.01 | 0.17 ± 0.02 |
| T2μ | 4.52 ± 0.04 | 4.49 ± 0.06 |
| Volume | 0.22 ± 0.02 | 0.18 ± 0.04 |
| Left SUB |  |  |
| T2σ | 0.15 ± 0.01 | 0.17 ± 0.02 |
| T2μ | 4.53 ± 0.04 | 4.50 ± 0.05 |
| Volume | 0.23 ± 0.03 | 0.18 ± 0.04 |
| Right EC |  |  |
| T2σ | 0.13 ± 0.01 | 0.16 ± 0.03 |
| T2μ | 4.58 ± 0.04 | 4.57 ± 0.04 |
| Volume | 0.26 ± 0.04 | 0.21 ± 0.06 |
| Left EC |  |  |
| T2σ | 0.14 ± 0.01 | 0.17 ± 0.02 |
| T2μ | 4.58 ± 0.04 | 4.57 ± 0.04 |
| Volume | 0.30 ± 0.04 | 0.26 ± 0.06 |
| Right BA35 |  |  |
| T2σ | 0.12 ± 0.01 | 0.14 ± 0.02 |
| T2μ | 4.65 ± 0.03 | 4.66 ± 0.07 |
| Volume | 0.24 ± 0.05 | 0.17 ± 0.05 |
| Left BA35 |  |  |
| T2σ | 0.12 ± 0.01 | 0.14 ± 0.02 |
| T2μ | 4.63 ± 0.04 | 4.62 ± 0.05 |
| Volume | 0.24 ± 0.05 | 0.20 ± 0.05 |

#### Results: T2 variation between subfields

##### T2 heterogeneity

The mixed model analysis revealed a significant main effect of subfield (F(4,576)=262, p<.0001) on T2 heterogeneity across different diagnoses in our mixed model analysis. The lowest T2 heterogeneity was observed in DG. CA and BA35 were significantly more heterogenous than DG (p_corr_ <.0001), but not different from each other (p_corr_ =1.00). EC was the next most heterogenous (vs CA123: p_corr_ <.0001), and SUB had the greatest T2 heterogeneity (vs EC: p_corr_ <.0001). We also observed a significant main effect of hemisphere (Left>Right, F(1,144)=14.1, p<.001) as well as an interaction between subfield and hemisphere (F(4,576)=7.16, p<.0001). Follow-up analysis of this interaction using two-way ANOVA with Bonferroni multiple comparisons test reveals significantly greater T2 heterogeneity selectively in DG in the left hemisphere compared to the right (p_corr_ =.0152) – but all other subfields did not differ between hemispheres.

##### Absolute T2

We also tested the relative absolute T2 between subfields. Our mixed model analysis revealed a substantial overall difference in T2μ between subfields (F(4,576)=974, p<.0001). DG had the highest T2μ, followed by CA, BA35, EC and finally SUB. All subfields were significantly different from each other at the p_corr_<.0001 level. We also observed a significant main effect of hemisphere (Right>Left, F(1,144)=7.72, p=.006), and a subfield*hemisphere interaction (F(4,576)=16.0, p<.0001). Follow-up analysis of this interaction using two-way ANOVA with Bonferroni multiple comparisons test reveals a significantly higher T2μ in left SUB (p_corr_ =.009) and a right PC (p_corr_ <.0001). A slightly increased T2μ was observed in right DG (p_corr_ =.077) and EC (p_corr_ =.082) compared to the left hemisphere, however neither of these were significant at the p=0.05 level after Bonferroni correction across subfields.

#### Discussion: T2 variation between subfields

We show several differences between absolute T2 in subfields of the MTL including a relatively prolonged T2 in the dentate gyrus compared to all other subfields, and a relatively short T2 in the subiculum. This could reflect different cell types and microstructural features of each subfield. For example, localised non-haem iron deposits (which decrease T2) have been shown to be more prevalent in the subiculum, and less so in DG (Antharam et al., 2012, Morris et al., 1994). Contrary to this, a review by Neely et al. (2019) shows how, at least in the rodent hippocampus, DG tends to have relatively high iron content, suggesting that iron may not be the primary driver of T2 differences between subfields. An alternative candidate for driving T2 changes could be demyelination. DG may be a subfield more susceptible to myelin loss in ageing, leading to the relatively high T2 seen in this subfield. This is supported by Radhakrishnan et al. (2020) who show that an MRI proxy for myelination in DG/CA3 is the best correlate of behaviour across all subfields. Furthermore, as DG is one of the last regions to develop myelin during development (Abrahám et al., 2010), it is supportive of the retrogenesis theory, which states that the brain degrades in an order in reverse to that in which it develops in AD (Reisberg et al., 2002). Even in a cohort of cognitively healthy older controls around 20% could be expected to harbour incipient AD pathology (Kern et al., 2018), explaining the relatively elevated T2 in DG.

The two known studies that have observed quantitative T2 in hippocampal subfields *in vitro* both found little difference between the subfields in absolute T2 (Huesgen et al., 1993, Antharam et al., 2012). This is in contrast to our findings, which may be due to *in vitro* vs *in vivo* methodological differences. T2 is also likely to vary throughout individual cell layers of allo- and neocortex given different density of cell bodies, proportions of glial cells and, as mentioned previously, densities of non-haem iron. Such differences in individual cortical layers within each subfield are lost here due to limitations in image resolution and contrast. Further exploration of these layers using ultra high-field MRI would be an interesting avenue of future research.

We have previously made the argument that any conclusions on the causes of changes in absolute T2 are difficult to make given the myriad different factors that affect T2 in opposing ways. We see that subfields with lower absolute T2 values have greater T2 heterogeneity. This suggests that the increases in T2 heterogeneity are brought about by a larger proportion of hypointense regions of T2 (T2-decreasing factors such as iron or myelin). However, when considering the effects of pathology, absolute T2 and T2 heterogeneity show very different patterns.

#### Path analysis - MCI

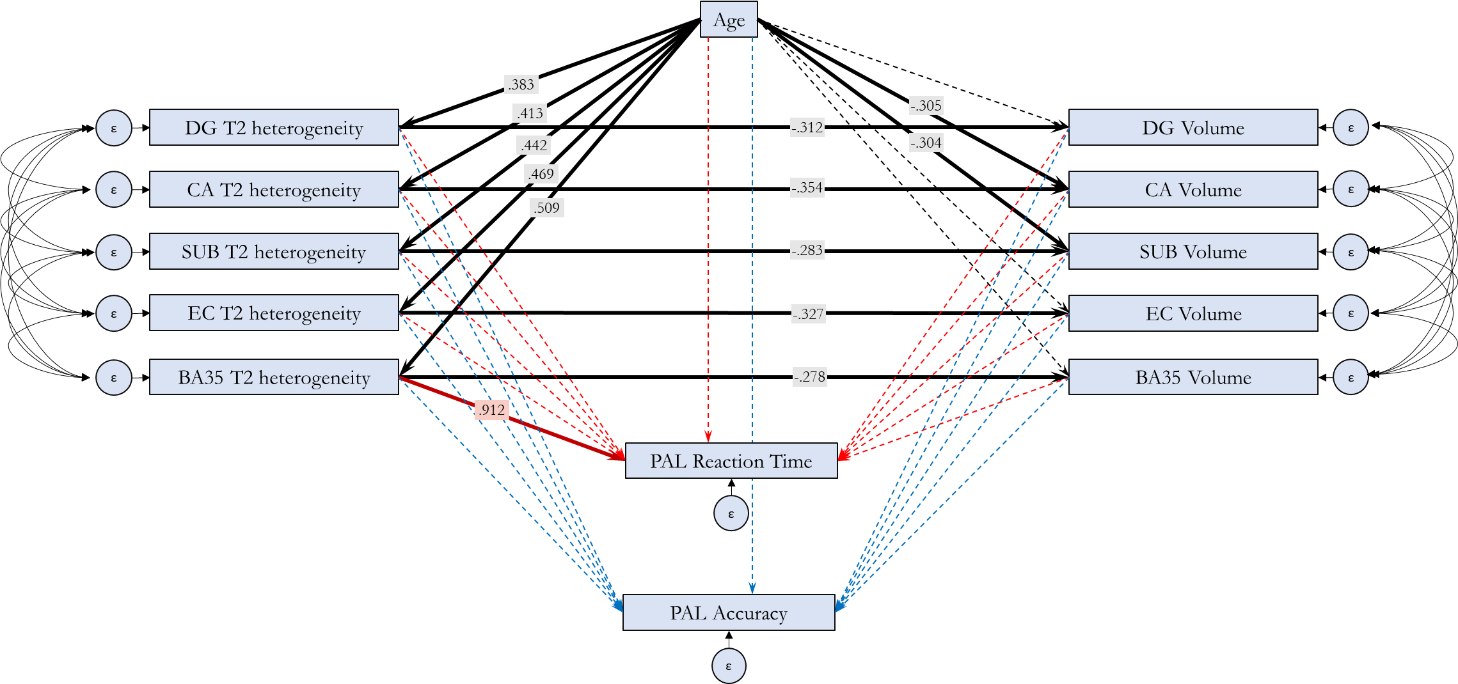

Figure 1 | Path analysis showing the relationship between Age, T2 heterogeneity, volume and memory in MTL subfields in people with MCI.

Bold arrows represent statistically significant relationships, with standardized B values indicated in overlaid boxes. Two models were run, each assessing one outcome measure of the PAL task. Black lines represent paths shared between the models. Unique paths to each model are shown in red (PAL reaction time as dependent variable) and blue (PAL Total Accuracy as dependent variable). Curves lines represent error term covariances defined in the model. All subfield volumes were normalised to ICV prior to entering into the model.

Model fit for MCI path analysis was poorer than that for healthy older controls (*χ^2^*=37.2, df = 20, p=.011; C_min_/DF=1.86; GFI=.888, AGFI=.564; CFI=.967; RMSEA=.143 ± 90% CI [.068-.214]; p_close_^=^.028).

Supplementary Table 4 | Covariance estimates for error terms within Path Analysis model for healthy older controls.
Values above the midline (yellow) represent covariance values between error terms on subfield volumes. Values below the midline (blue) represent covariance values between error terms on subfield T2σ (T2 heterogeneity).

|  | DG | | CA123 | | SUB | | EC | | PC | |
| --- | --- | --- | --- | --- | --- | --- | --- | --- | --- | --- |
|  | B ± SE | P | B ± SE | P | B ± SE | P | B ± SE | P | B ± SE | P |
| DG | - | - | .425 ± .081 | <.001 | .265 ± .075 | <.001 | .194 ± .070 | 0.006 | .160 ± .073 | 0.028 |
| CA123 | .757 ± .114 | <.001 | - | - | .411 ± .080 | <.001 | .268 ± .071 | <.001 | .166 ± .071 | 0.02 |
| SUB | .695 ± .109 | <.001 | .656 ± .105 | <.001 | - | - | .321 ± .075 | <.001 | .181 ± .073 | 0.013 |
| EC | .619 ± .102 | <.001 | .586 ± .099 | <.001 | .620 ± .101 | <.001 | - | - | .182 ± .070 | 0.01 |
| PC | .615 ± .101 | <.001 | .615 ± .100 | <.001 | .579 ± .097 | <.001 | .586 ± .096 | <.001 | - | - |

Supplementary Table 5 | Covariance estimates for error terms within Path Analysis model for people with MCI.
Values above the midline (yellow) represent covariance values between error terms on subfield volumes. Values below the midline (blue) represent covariance values between error terms on subfield T2σ (T2 heterogeneity).

|  | DG | | CA123 | | SUB | | EC | | PC | |
| --- | --- | --- | --- | --- | --- | --- | --- | --- | --- | --- |
|  | B ± SE | P | B ± SE | P | B ± SE | P | B ± SE | P | B ± SE | P |
| DG | - | - | .994 ± .231 | <.001 | .791 ± .221 | <.001 | .576 ± .203 | 0.005 | .500 ± .167 | 0.003 |
| CA123 | .937 ± .215 | <.001 | - | - | .966 ± .239 | <.001 | .743 ± .218 | <.001 | .643 ± .180 | <.001 |
| SUB | .836 ± .222 | <.001 | .844 ± .211 | <.001 | - | - | .942 ± .245 | <.001 | .711 ± .194 | <.001 |
| EC | .856 ± .279 | 0.002 | .987 ± .272 | <.001 | 1.33 ± .327 | <.001 | - | - | .638 ± .187 | <.001 |
| PC | .968 ± .245 | <.001 | .968 ± .233 | <.001 | 1.05 ± .259 | <.001 | 1.45 ± .355 | <.001 | - | - |
